# Dietary differences by job type and industry among workers in Japan during the COVID-19 pandemic: A cross-sectional study

**DOI:** 10.1101/2021.08.25.21262645

**Authors:** Rie Tanaka, Toshihide Sakuragi, Mayumi Tsuji, Seiichiro Tateishi, Ayako Hino, Akira Ogami, Masako Nagata, Shinya Matsuda, Yoshihisa Fujino, for the CORoNaWork Project

## Abstract

**Background:** The COVID-19 pandemic requires people to change their lifestyles. This study aimed to examine the differences in dietary behaviors during the pandemic across job types and industries.

**Methods:** A cross-sectional study was conducted using data from the Novel-coronavirus and Work Project. Job type and industry were classified into 3 and 22 groups, respectively. Dietary behaviors were assessed using self-reported questionnaires regarding eating breakfast, frequency of meals, and eating fast foods. Changes in eating breakfast during the pandemic were also evaluated. Logistic regression analysis nested in the workplace prefecture was carried out and adjusted for gender, age, body mass index, presence of family members, educational background, and household income.

**Results:** Workers involved in jobs that require communicating with people were more likely to skip breakfast (OR 1.17, 95%CI 1.10-1.24) and had a lower frequency of meals (OR 1.25, 95%CI 1.17-1.34) than workers engaged in desk work. Manual workers were more likely to eat fast food or meals (OR 1.10, 95%CI 1.03-1.17) than workers engaged in desk work. Workers engaged in newspaper, magazine, television, radio, advertising, and other mass media industries were more likely to skip breakfast (OR 2.43, 95%CI 1.82-3.24) and have a lower frequency of meals (OR 3.90, 95%CI 2.87-5.28) than workers in public offices and organizations.

**Conclusion:** The tendencies of dietary behaviors across job types/industries during the pandemic were shown, partially consistent with a trend reported before the pandemic. Further studies should clarify the causes of differences in dietary behaviors among workers.

## Introduction

The COVID-19 pandemic has changed people’s lifestyles (1). People were required to stay home to prevent the spread of COVID-19, which directly affected food accessibility and availability. Consequently, dietary changes have been reported worldwide. Previous studies have described changes in eating habits, such as eating a late-night snack or meal, having a fresh main meal, consuming less fast food (2), and more snacking (3).

Variation in dietary habits during the pandemic may depend on background characteristics, such as age, body mass index, and physical activities. A study in Spain reported that people with increased physical activity showed higher scores in the Healthy Mediterranean-Style dietary pattern (4). An Italian survey reported that the 18–30-year age group showed higher adherence to the Mediterranean diet than younger and older groups during lockdown (5). According to a study involving Polish adults, the tendency of eating and snacking seemed to be observed in individuals who were overweight and had obesity (6).

The present study focused on the working generation. During the pandemic, working styles may also have an impact on dietary behaviors. Variation in working style during the pandemic can differ according to job type and industry. Before the pandemic, several studies have shown workers’ diets according to their occupation (7-9). However, no similar studies have focused on the effects of the pandemic. Little is known about the dietary differences according to job type or industry during the pandemic.

In the COVID-19 crisis, a public health emergency, information on dietary differences between occupational groups can help promote healthy eating among working people. This study aimed to investigate the differences in dietary behaviors across job types and industrial groups during the pandemic.

## Methods

### Study design and participants

This cross-sectional study used data from the Collaborative Online Research on the Novel-coronavirus and Work (CORoNa Work) Project. This project is an Internet-based nationwide prospective cohort study designed to investigate the working environments and health status of workers during the COVID-19 pandemic in Japan. Data for the present study were obtained from the baseline survey, which was conducted during the third wave of the COVID-19 pandemic in Japan in December 2020. The study protocol has been described elsewhere (10) and was approved by the Ethics Committee of the University of Occupational and Environmental Health, Japan. Full-time workers, excluding healthcare workers or caregivers, were invited to participate in the survey. Participants (33,302) were selected based on stratification by gender, region, and job type. After excluding invalid responses, 27,036 participants were included in the study. Informed consent was obtained from the participants’ through the website.

### Measures

#### Job description and industry

Information on job type was assessed using a single question, “Please choose the response that is closest to your job description.” The answer categories were: mainly desk work (e.g., office work and computer work), mainly work involving communicating with people (e.g., customer service and sales), and manual work (e.g., work at a production site, manual labor, and nursing care). Industry information was also assessed using a single question, “Please choose the industry in which you work.” The 22 answer categories were as follows: energy, materials, and industrial machinery; food; beverages/tobacco products; pharmaceuticals/medical supplies; cosmetics/toiletries/sanitary products; fashion and accessories; precision machinery and office supplies; home appliances/AV equipment; automobiles and transportation equipment; household goods; hobby/sporting goods; real estate and housing equipment; information and communication; distribution and retail; finance/insurance; transportation and leisure; restaurant and other services; public offices and organizations; education, medical services, religion; newspapers, magazines, television, radio, advertising, and other mass media; market research; other.

#### Dietary behavior

The questionnaire regarding eating breakfast had five response categories, as follows: “6–7 days a week,” “4–5 days a week,” “2–3 days a week,” “less than 1 day a week,” and “hardly ever.” Questionnaires regarding the frequency of meals and eating only fast food or meals per day also had five response categories: “four or more times,” “three times,” “twice,” “once,” and “hardly ever.” Regarding eating breakfast, changes in frequency were also assessed using a single item, “We would like to ask you about the impact of the outbreak of COVID-19 on your social and living conditions. Please select the answer that best applies to the following items.” Response options were: “increased,” “stayed the same,” and “decreased.”

#### Sociodemographic factors

Participants responded to whether they lived with family members with “yes” or “no.” Household income was categorized into 3 groups: Less than 2,000,000 yen; 2,000,000–9,999,999 yen; 10,000,000 yen or more. Educational background was categorized into 3 groups: junior high school, high school; vocational school, technical c/junior college; university, graduate school.

### Data analysis

Descriptive analysis was performed using means and standard deviations for continuous variables and numbers and percentages for categorical variables. Logistic regression analysis was performed using Stata/IC 14.0 (StataCorp, College Station, TX, USA) to analyze the association between dietary behaviors and job type or industry. Mainly desk work and public offices and organizations were defined as the reference category for job type and industry group, respectively. Responses regarding dietary behaviors were divided into two categories: eating breakfast, 6–7 days and ≤5 days a week; frequency of meals, ≥3 times and ≤2 times a day; eating only fast food or meals, hardly ever and more than once a day. In this analysis, eating breakfast ≤5 days a week was defined as “Skipping breakfast,” eating ≤2 times a day was defined as “Lower frequency of meals,” and eating only fast food or meals more than once a day was defined as “Eating only fast food or meals.” All final models were adjusted for gender, age, body mass index, presence of family members, educational background, and household income. The odds ratio and 95% confidence interval (CI) were calculated. The level of statistical significance was set at *p* < 0.05.

## Results

### Participant characteristics

Table 1 shows the characteristics of the 27,036 participants. The mean age of the participants was 47.0 years (standard deviation, 10.5), and their body mass index was 22.4 (standard deviation, 4.7). Approximately half of the patients were men (51%), and 79% of the participants reported the presence of family members. Participants who reported a household income of 2,000,000–9,999,999 million yen were 78%. Nearly half of the participants reported that they had graduated from university. Regarding dietary behaviors, 63% reported eating breakfast 6–7 days a week, 75% reported eating meals 3 times a day, and 63% reported eating only fast food or meals. Most participants (93%) reported that their frequency of eating breakfast did not change during the pandemic period. Regarding job type, workers engaged in desk work were 50%, workers involved in jobs that require communicating with people were 26%, and manual workers were 25%. Among the 22 industries, education, medical services, and religion showed the highest percentage of participants (17%), except for the “other” group.

**Table 1.** Characteristics of the participants

|  | Mean | SD |
| --- | --- | --- |
| Age (year) | 47.0 | 10.5 |
| Body mass index | 22.4 | 3.7 |
| Gender | n | (%) |
| Male | 13,814 | (51.1) |
| Eat breakfast (days a week) |  |  |
| 6–7 | 17,078 | (63.2) |
| 5 or less than | 9,958 | (36.8) |
| Frequency of meals (times a day) |  |  |
| 3 or more than | 20,712 | (76.6) |
| 2 or less than | 6,324 | (23.4) |
| Eat only fast food or meals (times a day) |  |  |
| hardly ever | 17,110 | (63.3) |
| once or more | 9,926 | (36.7) |
| Days breakfast was eaten |  |  |
| increased | 1,161 | (4.3) |
| stayed the same | 25,092 | (92.8) |
| decreased | 783 | (2.9) |
| Family members |  |  |
| yes | 21,229 | (78.5) |
| Household income |  |  |
| Less than 2 million yen | 1,709 | (6.3) |
| 2 to 9.99 million yen | 21,077 | (78.0) |
| More than 10 million yen | 4,250 | (15.7) |
| Educational background |  |  |
| Junior high school, High school | 7,321 | (27.1) |
| Vocational school, technical college/junior college | 6,544 | (24.2) |
| University, Graduate school | 13,171 | (48.7) |
| Job type |  |  |
| Mainly desk work (e.g., office work, computer work) | 13,468 | (49.8) |
| Mainly work involving communicating with people (e.g., customer service, sales, etc.) | 6,927 | (25.6) |
| Manual work (e.g., work at a production site, manual labor, nursing care, etc.) | 6,641 | (24.6) |
| Industry |  |  |
| Energy, materials, industrial machinery | 980 | (3.6) |
| Food | 609 | (2.3) |
| Beverages/Tobacco products | 138 | (0.5) |
| Pharmaceuticals/Medical supplies | 396 | (1.5) |
| Cosmetics/Toiletries/Sanitary products | 159 | (0.6) |
| Fashion and Accessories | 312 | (1.2) |
| Precision Machinery and Office supplies | 464 | (1.7) |
| Home appliances/AV equipment | 481 | (1.8) |
| Automobiles and Transportation equipment | 911 | (3.4) |
| Household goods | 65 | (0.2) |
| Hobby/Sporting goods | 76 | (0.3) |
| Real estate and Housing equipment | 944 | (3.5) |
| Information and Communication | 1,331 | (4.9) |
| Distribution and Retail | 1,797 | (6.7) |
| Finance/Insurance | 1,226 | (4.5) |
| Transportation and Leisure | 671 | (2.5) |
| Restaurant and Other services | 1,319 | (4.9) |
| Public offices and Organizations | 1,891 | (7.0) |
| Education, medical services, religion | 4,500 | (16.6) |
| Newspapers, magazines, television, radio, advertising, and other mass media | 218 | (0.8) |
| Market research | 48 | (0.2) |
| Other | 8,500 | (31.4) |

### Dietary behaviors and job type/industry

Table 2 shows the association between skipping breakfast and the type of job/industry. Individuals who reported eating breakfast 6–7 days a week were as follows: 66% of desk workers, 61% of workers involved in jobs that require communicating with people, and 61% of manual workers. Compared with individuals engaged in desk work, workers involved in jobs that require communicating with people had higher chances of skipping breakfast (OR 1.17, 95%CI 1.10-1.24, *p < 0*.*001*). Individuals engaged in work related to newspapers, magazines, television, radio, advertising, and other mass media were more likely to skipp breakfast compared with individuals working in public offices and organizations (OR 2.43, 95%CI 1.82-3.24, *p < 0*.*001*). About half of them showed eating breakfast 5 or less than days a week.

**Table 2.** Association between skipping breakfast and job types/industries during the pandemic

|  | Breakfast (day a week) |  |  | Skipping breakfast |  |  |  |  |  |
| --- | --- | --- | --- | --- | --- | --- | --- | --- | --- |
|  | n | 6–7 | 5 or less<br>than | Crude |  |  | Adjusted* |  |  |
|  |  |  |  | OR | 95%CI |  | OR | 95%CI |  |
| Job type |  |  |  |  |  |  |  |  |  |
| Mainly desk work (e.g., office work, computer work) | 13,468 | 65.5 | 34.5 | Ref |  |  | Ref |  |  |
| Mainly work involving communicating with people (e.g., customer service, sales, etc.) | 6,927 | 60.7 | 39.3 | 1.23 | 1.16 | 1.31 | 1.17 | 1.10 | 1.24 |
| Manual work (e.g., work at a production site, manual labor, nursing care, etc.) | 6,641 | 61.0 | 39.0 | 1.22 | 1.14 | 1.29 | 1.04 | 0.98 | 1.11 |
| Industry |  |  |  |  |  |  |  |  |  |
| Public offices and Organizations | 980 | 71.5 | 28.5 | Ref |  |  | Ref |  |  |
| Energy, materials, industrial machinery | 609 | 66.1 | 33.9 | 1.29 | 1.09 | 1.52 | 1.21 | 1.03 | 1.44 |
| Food | 138 | 59.6 | 40.4 | 1.70 | 1.41 | 2.06 | 1.44 | 1.18 | 1.75 |
| Beverages/Tobacco products | 396 | 68.1 | 31.9 | 1.17 | 0.81 | 1.70 | 1.02 | 0.70 | 1.49 |
| Pharmaceuticals/Medical supplies | 159 | 65.4 | 34.6 | 1.33 | 1.05 | 1.67 | 1.22 | 0.96 | 1.54 |
| Cosmetics/Toiletries/Sanitary products | 312 | 53.5 | 46.5 | 2.18 | 1.57 | 3.03 | 1.93 | 1.38 | 2.70 |
| Fashion and Accessories | 464 | 58.0 | 42.0 | 1.82 | 1.42 | 2.32 | 1.52 | 1.18 | 1.96 |
| Precision Machinery and Office supplies | 481 | 66.0 | 34.1 | 1.30 | 1.04 | 1.61 | 1.19 | 0.95 | 1.48 |
| Home appliances/AV equipment | 911 | 65.3 | 34.7 | 1.33 | 1.08 | 1.65 | 1.24 | 1.00 | 1.54 |
| Automobiles and Transportation equipment | 65 | 60.8 | 39.2 | 1.62 | 1.37 | 1.91 | 1.39 | 1.17 | 1.64 |
| Household goods | 76 | 75.4 | 24.6 | 0.82 | 0.46 | 1.45 | 0.76 | 0.42 | 1.35 |
| Hobby/Sporting goods | 944 | 67.1 | 32.9 | 1.23 | 0.75 | 2.00 | 1.19 | 0.72 | 1.96 |
| Real estate and Housing equipment | 1,331 | 60.4 | 39.6 | 1.65 | 1.40 | 1.94 | 1.60 | 1.35 | 1.89 |
| Information and Communication | 1,797 | 65.2 | 34.8 | 1.34 | 1.15 | 1.56 | 1.27 | 1.09 | 1.48 |
| Distribution and Retail | 1,226 | 59.4 | 40.6 | 1.71 | 1.49 | 1.96 | 1.57 | 1.36 | 1.80 |
| Finance/Insurance | 671 | 63.4 | 36.6 | 1.45 | 1.24 | 1.69 | 1.40 | 1.20 | 1.64 |
| Transportation and Leisure | 1,319 | 55.9 | 44.1 | 1.98 | 1.65 | 2.38 | 1.72 | 1.43 | 2.08 |
| Restaurant and Other services | 1,891 | 56.7 | 43.3 | 1.91 | 1.65 | 2.22 | 1.68 | 1.45 | 1.96 |
| Education, medical services, religion | 4,500 | 66.2 | 33.8 | 1.28 | 1.14 | 1.44 | 1.21 | 1.07 | 1.36 |
| Newspapers, magazines, television, radio, advertising, and other mass media | 218 | 49.1 | 50.9 | 2.60 | 1.96 | 3.46 | 2.43 | 1.82 | 3.24 |
| Market research | 48 | 56.3 | 43.8 | 1.95 | 1.09 | 3.48 | 1.73 | 0.96 | 3.13 |
| Other | 8,500 | 62.4 | 37.6 | 1.51 | 1.36 | 1.69 | 1.40 | 1.25 | 1.56 |

\* Adjusted for gender, age, bmi, family member, educational background, and household income

Table 3 shows the association between lower frequency of meals and the type of job/industry. Individuals who reported eating meals ≥3 times a day were as follows: 78% of desk workers, 74% of workers involved in jobs that require communicating with people, and 76% of manual workers. Compared with individuals engaged in desk work, workers involved in jobs that require communicating with people had higher chances of lower frequency of meals (OR 1.25, 95%CI 1.17-1.34, *p < 0*.*001*). Individuals engaged in work related to newspapers, magazines, television, radio, advertising, and other mass media were more likely to have lower frequency of meals compared with individuals working in public offices and organizations (OR 3.90, 95%CI 2.87-5.28, *p < 0*.*001*). Among them, 40% showed eating meals 2 or less times a day.

**Table 3.**
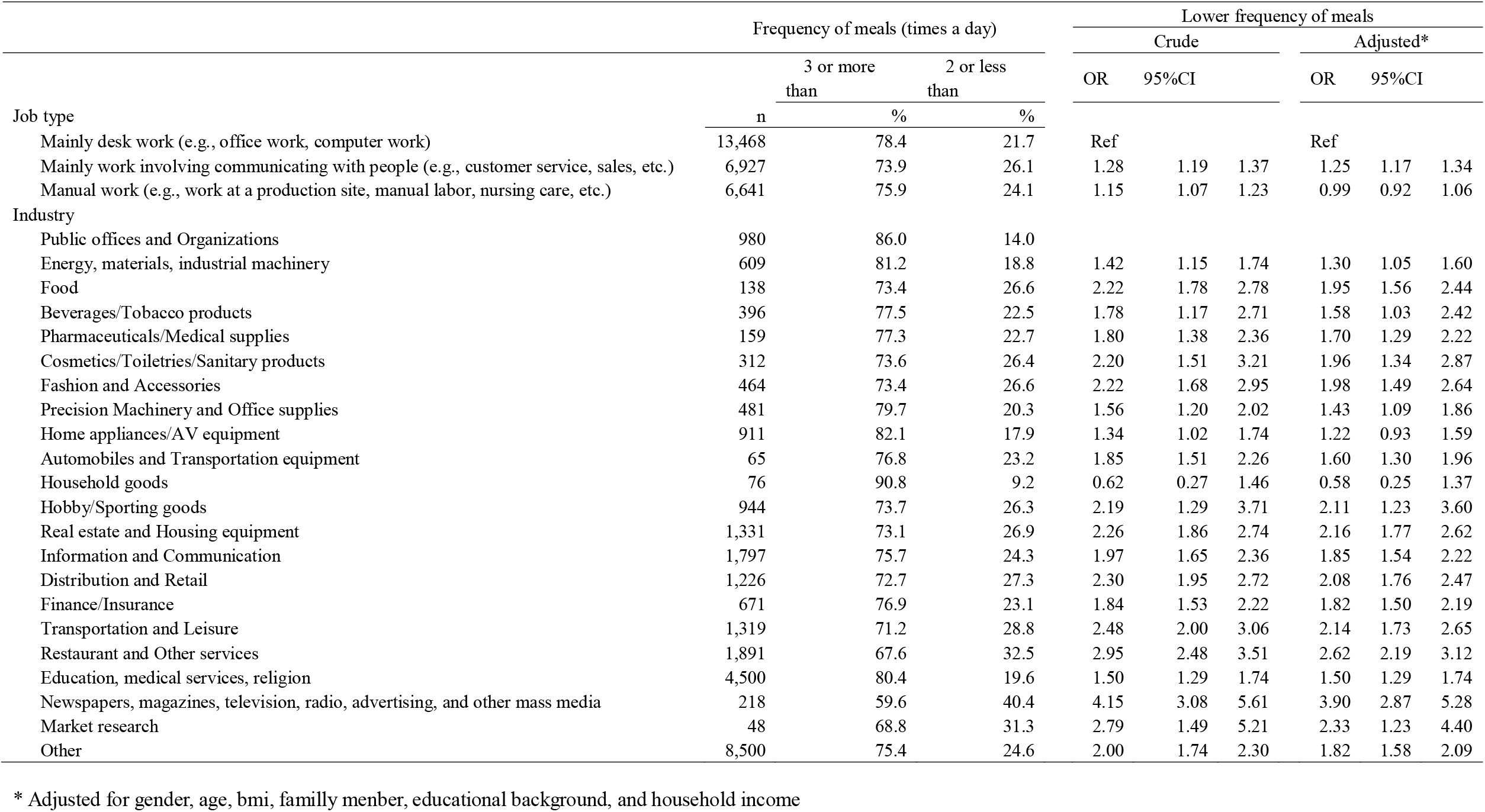
Association between lower frequency of meals and job types/industries during the pandemic

Table 4 shows the association between eating only fast food and the type of job/industry. Individuals who reported eating only fast food or meals were as follows: 65% of desk workers, 63% of workers involved in jobs that require communicating with people, and 61% of manual workers. Compared with individuals engaged in mainly desk work, manual workers were more likely to eat only fast food or meals (OR 1.10, 95%CI 1.03-1.17, *p = 0*.*005*).

**Table 4.** Association between eating only fast food and job types/industries during the pandemic

|  | Eat only fast food or meals<br>(times a day) |  |  | Eat only fast food or meals |  |  |  |  |  |
| --- | --- | --- | --- | --- | --- | --- | --- | --- | --- |
|  | n | hardly<br>ever | once or<br>more | Crude |  |  | Adjusted* |  |  |
|  |  | % | % | OR | 95%CI |  | OR | 95%CI |  |
| Job type |  |  |  |  |  |  |  |  |  |
| Mainly desk work (e.g., office work, computer work) | 13,468 | 64.7 | 35.3 | Ref |  |  | Ref |  |  |
| Mainly work involving communicating with people (e.g., customer service, sales, etc.) | 6,927 | 62.8 | 37.2 | 1.09 | 1.02 | 1.15 | 1.05 | 0.99 | 1.12 |
| Manual work (e.g., work at a production site, manual labor, nursing care, etc.) | 6,641 | 60.9 | 39.1 | 1.17 | 1.11 | 1.25 | 1.10 | 1.03 | 1.17 |
| Industry |  |  |  |  |  |  |  |  |  |
| Public offices and Organizations | 980 | 67.7 | 32.3 | Ref |  |  | Ref |  |  |
| Energy, materials, industrial machinery | 609 | 61.3 | 38.7 | 1.32 | 1.13 | 1.55 | 1.30 | 1.10 | 1.52 |
| Food | 138 | 61.6 | 38.4 | 1.31 | 1.08 | 1.58 | 1.19 | 0.98 | 1.44 |
| Beverages/Tobacco products | 396 | 60.9 | 39.1 | 1.35 | 0.94 | 1.92 | 1.26 | 0.88 | 1.80 |
| Pharmaceuticals/Medical supplies | 159 | 60.6 | 39.4 | 1.36 | 1.09 | 1.70 | 1.30 | 1.04 | 1.62 |
| Cosmetics/Toiletries/Sanitary products | 312 | 61.0 | 39.0 | 1.34 | 0.96 | 1.87 | 1.22 | 0.87 | 1.71 |
| Fashion and Accessories | 464 | 65.1 | 34.9 | 1.12 | 0.87 | 1.45 | 1.00 | 0.77 | 1.29 |
| Precision Machinery and Office supplies | 481 | 63.6 | 36.4 | 1.20 | 0.97 | 1.48 | 1.15 | 0.92 | 1.42 |
| Home appliances/AV equipment | 911 | 63.4 | 36.6 | 1.21 | 0.98 | 1.49 | 1.17 | 0.95 | 1.45 |
| Automobiles and Transportation equipment | 65 | 61.3 | 38.8 | 1.33 | 1.12 | 1.56 | 1.24 | 1.05 | 1.46 |
| Household goods | 76 | 56.9 | 43.1 | 1.59 | 0.96 | 2.61 | 1.51 | 0.91 | 2.49 |
| Hobby/Sporting goods | 944 | 60.5 | 39.5 | 1.37 | 0.85 | 2.19 | 1.33 | 0.83 | 2.14 |
| Real estate and Housing equipment | 1,331 | 62.7 | 37.3 | 1.25 | 1.06 | 1.47 | 1.22 | 1.04 | 1.44 |
| Information and Communication | 1,797 | 62.7 | 37.3 | 1.24 | 1.07 | 1.44 | 1.20 | 1.04 | 1.39 |
| Distribution and Retail | 1,226 | 59.9 | 40.1 | 1.40 | 1.23 | 1.61 | 1.33 | 1.16 | 1.52 |
| Finance/Insurance | 671 | 64.1 | 35.9 | 1.17 | 1.01 | 1.36 | 1.14 | 0.98 | 1.33 |
| Transportation and Leisure | 1,319 | 61.3 | 38.8 | 1.33 | 1.10 | 1.59 | 1.22 | 1.02 | 1.47 |
| Restaurant and Other services | 1,891 | 62.7 | 37.3 | 1.25 | 1.08 | 1.44 | 1.14 | 0.98 | 1.33 |
| Education, medical services, religion | 4,500 | 65.0 | 35.0 | 1.13 | 1.01 | 1.26 | 1.07 | 0.95 | 1.20 |
| Newspapers, magazines, television, radio, advertising, and other mass media | 218 | 64.7 | 35.3 | 1.14 | 0.85 | 1.54 | 1.08 | 0.80 | 1.45 |
| Market research | 48 | 68.8 | 31.3 | 0.95 | 0.51 | 1.77 | 0.86 | 0.46 | 1.61 |
| Other | 8,500 | 63.1 | 36.9 | 1.23 | 1.10 | 1.36 | 1.17 | 1.05 | 1.30 |

\* Adjusted for gender, age, bmi, family member, educational background, and household income

Figure 1 showed changes in breakfast eating according to job types/industries. Across all job types, more than 90% of workers reported that they did not change the frequency of breakfast eating. Across all industries, 87%–96% maintained the same frequency of breakfast.

**Figure 1.**
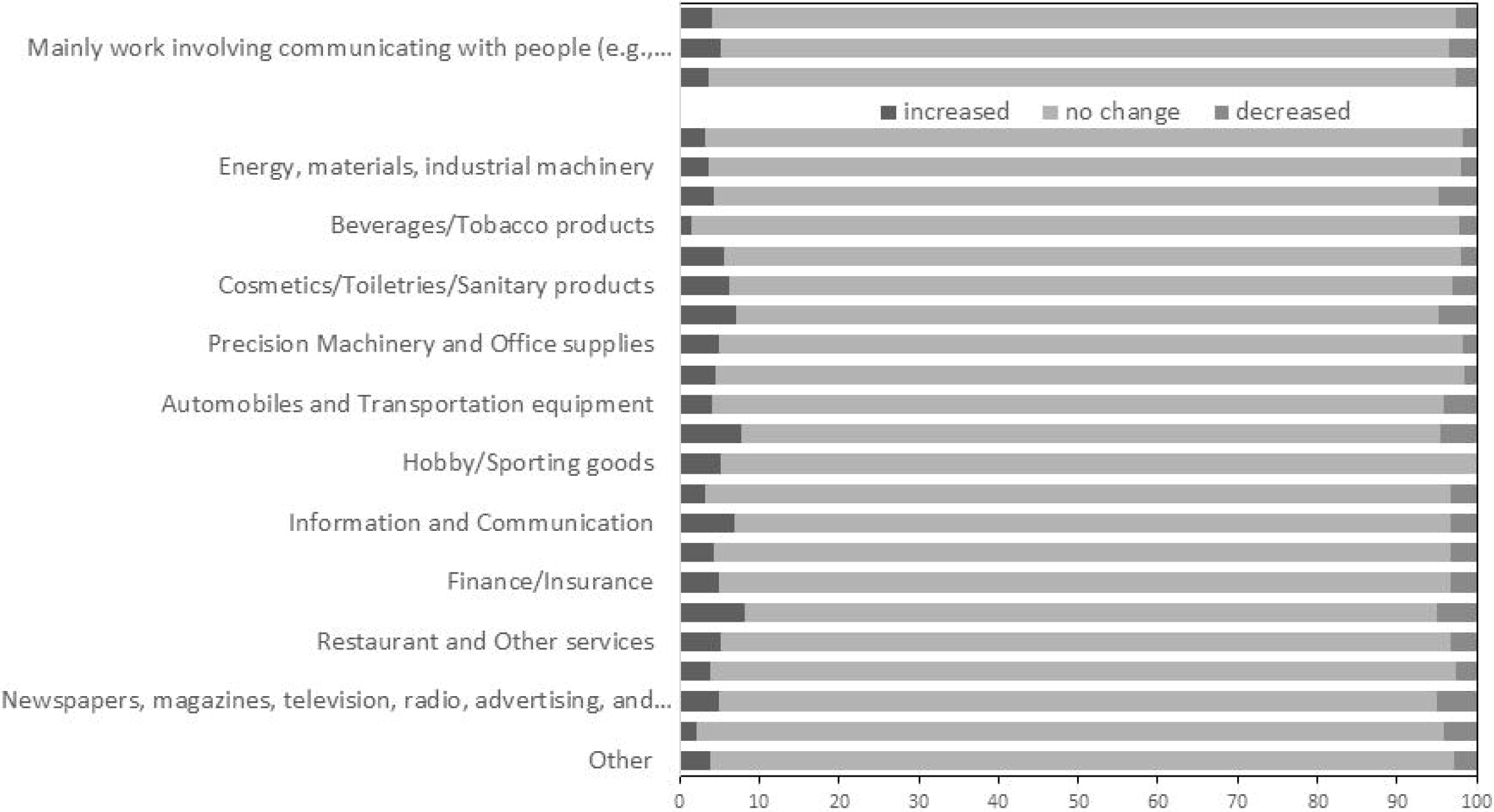
Changes in breakfast eating according to job types/industries

## Discussion

This study found differences in dietary behaviors according to the type of job or industry during the COVID-19 pandemic in Japan. Compared with mainly desk workers, workers involved in jobs that require communicating with people (e.g., customer service and sales) were more likely to skip breakfast or other meals, and those involved in manual work (e.g., work at a production site, manual labor, and nursing care) were more likely to eat fast food. Compared with individuals working in public offices and organizations, many workers skipped breakfast or other meals. Workers engaged in industries involving newspapers, magazines, television, radio, advertising, and other mass media showed a lower meal frequency.

This study describes dietary differences according to job type during the pandemic. The tendency to skip breakfast and have lower frequency of meals among workers involved in jobs that require communicating with people compared with mainly desk workers was consistent with a previous study conducted before the pandemic. Merchandise sales workers and other service workers showed higher likelihoods of skipping breakfast compared to management government officials (9). The tendency of manual workers to eat fast food compared with desk workers was also consistent with the same study; motor vehicle drivers showed a higher likelihood of eating instant food compared to management government officials (9). The characteristics of dietary behaviors among workers engaged in such jobs should be considered in the field of occupational health, regardless of the COVID-19 outbreak.

The study also clarified dietary differences between industries. In particular, workers in newspaper, magazine, television, radio, advertising, and other mass media industries showed the highest percentage of skipping breakfast and had lower frequency of meals. Of the 218 participants belonging to this industry, 51% reported that they ate breakfast ≤5 days a week, and 40% reported that they ate meals ≤2 times a day. To our knowledge, no studies have investigated dietary behaviors among workers engaged in industries involving newspapers, magazines, television, radio, advertising, and other mass media, regardless of period, including before the pandemic.

This finding may be partially explained by their working conditions, including several work-related factors such as shifts and work hours. For example, one previous study reported that longer work hours were associated with a lower likelihood of a healthier diet (11); that is, work hours were shown to be associated with dietary habits. In 2020, a relatively small decrease in work hours was shown in industries related to information and communications, including newspapers, magazines, television, radio, advertising, and other mass media. In contrast, a large decrease in work hours was shown in service workers (12). With the association between work hours and dietary behaviors during the pandemic, there would have been relatively small changes in dietary behaviors among workers in industries related to information and communications. From the perspective of occupational health, understanding dietary tendencies regardless of before or during the pandemic among workers engaged in such industries is critical.

In this study, most participants reported that they did not change their breakfast eating habits. A previous review summarized papers reporting changes in dietary habits before and during the pandemic: snacking, meal number and frequency, fresh produce and home cooking, comfort food, and alcohol consumption (13). However, regarding the frequency of breakfast eating, little evidence is available; one study in Kuwait showed no significant change in skipping breakfast (2). In our study, although most of the workers who participated showed no change in the frequency of breakfast eating, there were several percentages of workers who reported increased or decreased frequency of breakfast eating. Differences in dietary behavioral changes during the pandemic may vary according to background characteristics, including gender, age, physical activity, and work-related factors. Further research regarding these workers with an increased or decreased chance of breakfast eating may help improve the dietary approach considering occupational health.

In summary, this study clarified dietary behaviors such as breakfast eating and fast-food eating according to job type during the COVID-19 pandemic in Japan; this was consistent with a study reported before the pandemic. Compared with public officers and organizations, most industries, especially newspapers, magazines, television, radio, advertising, and other mass media industries, showed a higher likelihood of lower frequency of meals, which was a novel finding of this study.

## Limitations

This study has several limitations. First, external validity and selection bias should be acknowledged. This analysis was based on data from an Internet survey, in which participants may not represent the national population. To reduce the effect of bias, the study sampled participants according to gender, region, and job type. Second, this study used the classification of 3 job types and 22 industrial groups. The industrial group includes various occupations. One large occupational group comprises various small occupational groups, among which there are differences in dietary intake (8). Similarly, one large industrial group contains various occupational groups, which may show different dietary behaviors; for example, according to the Japan Standard Industrial Classification, Rev.13, television broadcasting, radio broadcasting, newspaper publishers, commercial art, and graphic design belong to Information and Communications (14). Collectively, information on both occupation and industry requires a discussion of workers’ dietary habits.

Furthermore, an occupation is occasionally related to desk work, service work, and manual work. To interpret our findings, it is necessary to have detailed information regarding the relationships among industry, occupation, and job type. Third, dietary behaviors were assessed using a self-reported questionnaire, which may have been influenced by social desirability bias. The potential of over- or under-estimation should be considered when evaluating the results of studies examining dietary intake. Finally, the potential confounding factors should be addressed. Our final models were adjusted for several socioeconomic factors, which are well known to be associated with dietary habits (15), whereas the OR changed after the adjustment. Additionally, another potential confounding factor should be considered, for example, information on shift work, which has been shown to be associated with dietary habits (11, 16-22). To examine a more detailed relationship between work and diet, future studies should consider work-related factors, which may be associated with dietary behaviors.

## Conclusion

This study observed the tendency of skipping meals among service workers and eating fast food among manual workers, compared to desk workers. Workers engaged in industries related to newspapers, magazines, television, radio, advertising, and other mass media were more likely to skip meals than workers in public offices and organizations. Our findings may be useful to promote a healthy diet according to occupation.

## Data Availability

Data not available due to ethical restrictions

## Acknowledgements

This study was supported and partly funded by the research grant from the University of Occupational and Environmental Health, Japan (no grant number); Japanese Ministry of Health, Labour and Welfare (H30-josei-ippan-002, H30-roudou-ippan-007, 19JA1004, 20JA1006, 210301-1, and 20HB1004); Anshin Zaidan (no grant number), the Collabo-Health Study Group (no grant number), and Hitachi Systems, Ltd. (no grant number) and scholarship donations from Chugai Pharmaceutical Co., Ltd. (no grant number)

The current members of the CORoNaWork Project, in alphabetical order, are as follows: Dr. Yoshihisa Fujino (present chairperson of the study group), Dr. Akira Ogami, Dr. Arisa Harada, Dr. Ayako Hino, Dr. Hajime Ando, Dr. Hisashi Eguchi, Dr. Kazunori Ikegami, Dr. Kei Tokutsu, Dr. Keiji Muramatsu, Dr. Koji Mori, Dr. Kosuke Mafune, Dr. Kyoko Kitagawa, Dr. Masako Nagata, Dr. Mayumi Tsuji, Ms. Ning Liu, Dr. Rie Tanaka, Dr. Ryutaro Matsugaki, Dr. Seiichiro Tateishi, Dr. Shinya Matsuda, Dr. Tomohiro Ishimaru, and Dr. Tomohisa Nagata. All members are affiliated with the University of Occupational and Environmental Health, Japan.

## Notes

### Competing Interest Statement

The authors have declared no competing interest.

### Author Declarations

Ethics Committee of the University of Occupational and Environmental Health, Japan

